# Population-Wide Assessment of Heart Rhythm and Physical Activity from 14-Day Recordings: The UK Biobank Cardiac Monitoring Study

**DOI:** 10.64898/2026.02.23.26346310

**Authors:** Stefan van Duijvenboden, Ahmed El-Medany, Hesham Aggour, Michele Orini, Wenjia Bai, John E J Gallacher, Jemma C Hopewell, Stephen Bell, Fu Siong Ng, Aiden Doherty, Barbara Casadei

**Affiliations:** Nuffield Department of Population Health, University of Oxford, Oxford, UK; National Heart and Lung Institute, Imperial College London, London, UK; School of Biomedical Engineering & Imaging Sciences, KCL, London, UK; Department of Computing, Department of Brain Sciences and Data Science Institute, Imperial College London, London, UK; Department of Psychiatry, University of Oxford, Oxford, UK; UK Biobank, Adswood, Stockport, Cheshire, UK; Department of Cardiology, Imperial College Healthcare NHS Trust, London, UK; Department of Cardiology, Chelsea and Westminster Hospital NHS Foundation Trust, London, UK

## Abstract

**Background:** Long-term electrocardiogram (ECG) monitoring with wearable devices enables large-scale characterisation of cardiac rhythms, but population-based evidence remains limited. The UK Biobank Cardiac Monitoring Study integrates 14-day patch-based ECG monitoring with accelerometry and detailed phenotypic and lifestyle data. Here, we report the acquisition protocol, data processing, and initial findings from 27,658 participants.

**Methods:** Participants in the UK Biobank imaging study were invited to undergo 14-day cardiac monitoring using a Zio XT (pilot phase; 2015–18) or BodyGuardian MINI (main phase; 2019– ongoing) monitor. ECGs were analysed by certified technicians and automated algorithms to identify atrial, ventricular, and conduction arrhythmias. In parallel, beat-to-beat RR intervals were derived using in-house algorithms, and physical activity from calibrated triaxial accelerometer data. Analyses assessed wear time, arrhythmia prevalence, circadian patterns, and repeatability.

**Findings:** In total, 27,658 participants (mean age 71 years; 49.9% women) were analysed, including 7,795 from the pilot phase and 21,141 from the main phase; 1,353 (4.9%) had repeat recordings. In the main phase, median wear time was 13.2 days (IQR 11.9–13.9), and undiagnosed atrial fibrillation (AF) was detected more frequently in men than women (3.2% vs 1.7%; p<0.001); 68% was paroxysmal, with 27.4% detected during week two. Ventricular tachycardia occurred in 12.1% (8.4% in women), with sustained episodes rare (0.4%) but observed. Arrhythmia timing varied markedly with activity, with AF peaking during nocturnal inactivity and ventricular ectopy increasing during activity, peaking at midday. Repeat assessments showed strong reproducibility of diurnal heart rate and activity profiles, with more modest arrhythmia consistency.

**Interpretation:** Extended ECG monitoring enables detection of subclinical arrhythmias and long-term physiological rhythms in older adults. Linkage to imaging, multi-omics, and clinical outcomes in UK Biobank will enable unprecedented evaluation of the natural history of asymptomatic rhythm disturbances and their impact on brain health.

**Funding:** British Heart Foundation and Wellcome Trust.

## Introduction

Wearable sensors have revolutionised cardiovascular monitoring by enabling continuous, long-term, and unobtrusive acquisition of high-quality data at scale^1^. One area of particular relevance is long-term ambulatory electrocardiogram (ECG) monitoring, which has been shown to improve the detection of arrhythmias substantially^2–4^, particularly atrial fibrillation (AF), a common arrhythmia that is frequently clinically silent yet associated with a potentially preventable increased risk of stroke, heart failure, and mortality^5^. Beyond arrhythmia detection, prolonged ECG and heart rate monitoring during daily activities offers valuable insights into circadian variation^6–8^, autonomic nervous system function^6,7^, and cardiorespiratory fitness^9^. These physiological patterns are significantly influenced by age^10^ and altered in the presence of cardiovascular^6,11–13^ and neurodegenerative disease^14–16^. Accordingly, wearable technologies are increasingly integrated into both clinical and large-scale epidemiological studies^3,17–19^.

Despite advances in sensor technology, significant gaps remain in fully leveraging this information for disease prediction and for identifying underlying causative mechanisms in large-scale prospective studies. For instance, insights into the clinical relevance, triggers, and substrates of device-detected subclinical arrhythmias remain limited by small sample size, narrow scope of arrhythmia types studied, and reliance on patient-based cohorts^2^. Furthermore, the scarcity of large-scale, prospective datasets that integrate wearable sensor data with complementary clinical measurements, longitudinal health outcomes, and multi-omics and imaging information limits investigation into the prediction and clinical significance of subclinical heart rhythm disturbances and disrupted physiological variations, such as circadian rhythms and sleep patterns^8^.

Here, we describe the acquisition protocol, data processing pipelines, and quality metrics for the UK Biobank Cardiac Monitoring Study, and present initial findings from approximately 28,000 paired ECG and accelerometer recordings, including arrhythmia prevalence and circadian patterns of heart rate and activity. Once recruitment is completed, these datasets will be linked to the wealth of genetic and phenotypic information that is available in UK Biobank participants^20^, enabling an unprecedented evaluation of the causes and natural history of asymptomatic rhythm disturbances and their impact on brain health.

## Methods

### Study population & data collection

UK Biobank (http://www.ukbiobank.ac.uk) is a prospective cohort study of ∼500,000 adults (40–69 years) recruited between 2006 and 2010 in 22 centres across the UK. The study was approved by the North-West Research Ethics Committee (06/MRE08/65), with long-term health follow-up via health records^20^. An imaging substudy (2015-2025) added head, heart and body MRI scans, carotid artery ultrasound examination, a DEXA scan and repeated cognitive assessment in 100,000 participants from the existing cohort^21^. A subset of these participants was invited to wear a patch cardiac monitor for 14 days, either applied by a member of staff at the imaging centre or self-applied within 6 weeks.

Cardiac monitoring was conducted in two phases and is nearing completion (Figure 1). A pilot phase (May 2015–December 2018) evaluated feasibility using Zio XT patch ECG monitors (iRhythm Technologies), initially in 200 UK Biobank imaging participants irrespective of age or atrial fibrillation (AF) status, followed by a larger pilot (N=7,795) in participants aged ≥60 years without prior AF or anticoagulant use. The Zio XT device recorded continuous single-lead ECG (∼200 Hz) and low-frequency triaxial accelerometry (∼1.56 Hz). The main study phase began in August 2019 and used BodyGuardian MINI devices (Preventice Solutions; now Boston Scientific), providing higher-resolution ECG (250 Hz) and accelerometry (25 Hz). The age threshold was set at ≥65 years, with temporary inclusion of participants aged ≥60 years during the COVID-19 pandemic, and inclusion of individuals with prior AF or anticoagulant use from May 2022. Eligibility screening and acceptance rates were systematically recorded, except for the first 521 deployments in the pilot phase (up to October 2016). In both study phases, devices were worn for 14 days and returned to the manufacturer, which provided UK Biobank with raw ECG and accelerometry data and arrhythmia annotations. A subset of participants undergoing repeat imaging were invited to complete a second monitoring period.

**Figure 1.**
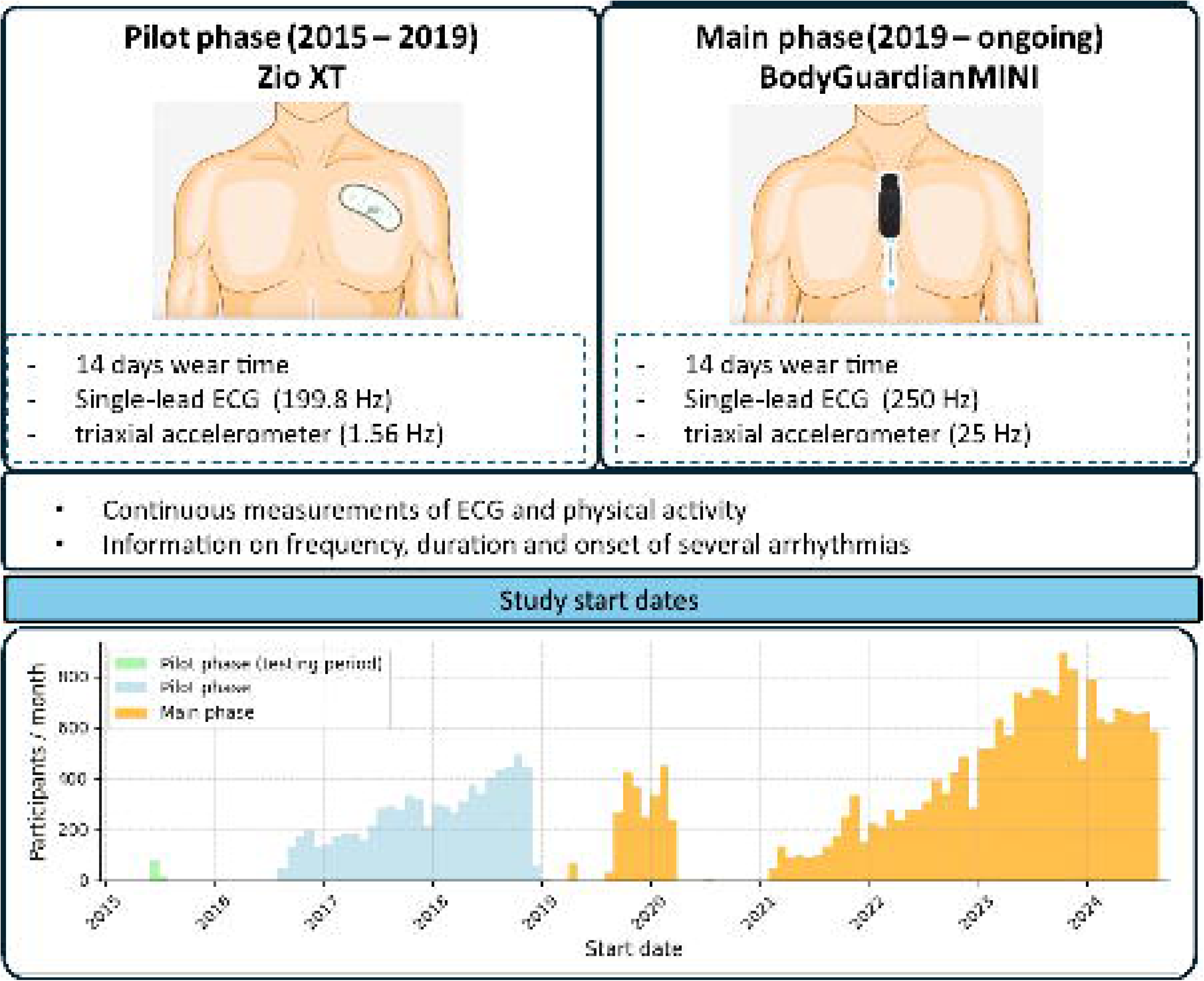
Overview of the UK Biobank cardiac monitor study. Data collection has been conducted over 2 phases and is currently ongoing.

### ECG data analysis & arrhythmia diagnoses

All ECG data from the main study phase (BodyGuardian MINI monitors) were processed by Preventice’s analysis partner (MDT) for quality control and arrhythmia assessment using automated algorithms (ECGLAB, Biomedical Instruments Co. Ltd.) alongside expert manual review. Certified MDT technicians used the BI CardioClient/ ECGLAB Holter system, combining automated QRS detection with manual validation and full-record review to ensure accurate beat labelling prior to summary metric generation. All analyses were conducted using the same software version, with final quality control checks completed by a senior technician. A list of analysed arrhythmias and their definitions is provided in Table 1. AF was classified as previously unreported if episodes >30 s were detected in participants who self-reported no prior AF at screening. This definition applied only to AF, as prior AF status was part of study eligibility, and AF detected during monitoring is therefore referred to as “new AF”, consistent with contemporary definitions of subclinical AF^22^. Supraventricular tachycardia (SVT) episodes of >30s in duration were counted.

**Table 1.** Definitions of cardiac arrhythmias analysed in the main study phase (2019-onwards).

| Arrhythmia event | Definition |
| --- | --- |
| AFib (Atrial Fibrillation) | Irregular polymorphic atrial activity with atrioventricular deficit (> 30s) |
| AFlut (Atrial Flutter) | Fast regular monomorphic atrial activity with atrioventricular deficit (> 30s) |
| SVT (Supraventricular Tachycardia) | Regular or slightly irregular non-sinus tachycardia with a narrow QRS complex (episode >30s) |
| VT (Ventricular Tachycardia) | Regular or slightly irregular tachycardia with a wide QRS complex with heart rate > 100 bpm (>3 QRS complexes) |
| IVR (Idioventricular Rhythm) | Regular or irregular rhythm with a wide QRS complex with rate < 100 bpm |
| Pause | R-R interval > 2 seconds |
| VF (Ventricular Fibrillation) | Irregular tachycardia with a wide polymorphic QRS complex |
| AV (Atrioventricular) Block I | Prolonged transmission from the atrial impulse to the ventricles with PR interval > 200ms |
| AV Block II | Intermittent failure of atrial impulses to conduct to the ventricles, due to a delay or interruption within the AV node or His–Purkinje system. This results in some P waves not being followed by a QRS complex |
| AV Block III | Complete atrioventricular block with dissociation between the atria and the ventricles |
| Isolated VE (Ventricular Ectopy) | Premature extra heartbeat originating from the ventricles, occurring earlier than the next expected normal beat |
| Isolated SVE (Supraventricular Ectopy) | Premature extra heartbeat originating from above the ventricles, occurring earlier than the next expected normal beat |
| VE Couplets / Triplets | Two or 3 VEs in a row |
| SVE Couplets / Triplets | Two or 3 SVEs in a row |
| Ventricular Bigeminy / Trigeminy | ≥Three cycles where every second (bigeminy) or third (trigeminy) beat is a ventricular ectopic |

For AF, atrial flutter, SVT, ventricular tachycardia (VT) and idioventricular rhythm, episode onset and durations were reported by MDT; remaining arrhythmias were reported as participant-level summaries. We analysed episode onset times and hourly summary data to assess diurnal variations in time spent in AF. We also calculated mean hourly frequencies of supraventricular ectopy (SVE) and ventricular ectopy (VE) by first calculating per-participant median ectopic counts per hour across the 14-day recording period and then averaging these medians across participants. Diurnal variation analyses were restricted to participants with ≥3 days of analysable ECG data.

ECG recordings derived during the pilot phase were analysed for a comparable set of arrhythmias (Supplemental Table 3) by iRhythm Technologies, using a proprietary algorithm approved by the US Food and Drug Administration, combined with trained cardiac technician review. However, data were provided in summary format only (e.g., without episode onset times or durations).

### Processing of ECG data

Because manufacturer-processed data did not include beat-level information, beat-to-beat RR intervals and ECG quality were derived using a deep learning–based QRS detection algorithm with predefined exclusions for non-wear, noise, and unstable signal (see Supplementary Methods). To quantify heart rate and its distribution, we computed median RR intervals across 10-second non-overlapping ECG epochs. Non-wear and unanalysable segments were imputed using the mean RR interval for the corresponding minute across the remaining days^23^. Participants were excluded if they had <3 days (72 hours) of analysable data or lacked coverage in each one-hour period of the 24-hour cycle.

### Processing of accelerometer data

To account for differences in sampling frequency and resolution between devices, accelerometer data were harmonised using a device-specific preprocessing pipeline. Recordings with >1% clipped values (beyond ±2g for Zio XT or ±4g for BodyGuardian MINI) were excluded. Signals were calibrated using the *Actipy* Python package (version 3.4.2)^24^ to correct for sensor bias^25^, after which movement-related acceleration was derived as the Euclidean norm of the calibrated axes. A median filter (4.8s window) was applied to attenuate remaining low-frequency baseline shifts before truncating negative values. Data were summarised in 1-minute epochs and non-wear periods, identified from ECG data, were imputed using the corresponding minute on other days, consistent with previous work^23^.

### Statistical analysis

Descriptive statistics were used to report device wear time, compliance in days, and accelerometer-measured physical activity (mg). Age groups were categorised into ≤70 and >70 years. Age and sex differences were assessed using the Wilcoxon–Mann–Whitney test for continuous data and Pearson’s chi-squared or Fisher’s exact test for categorical data. Correlation, morphological correlation, and intra-class correlation coefficients were used to assess agreement and repeatability of heart rate and acceleration measures across repeated recordings.

## Results

During the pilot phase, 7,869 of 12,981 invited participants (61%) consented, together with 521 recruited before eligibility was systematically recorded (Figure 2; Supplementary Table 2). In the main phase, 22,858 of 39,123 invited participants (58%) consented (Figure 2; Supplementary Table 3). After exclusions for missing data (Figure 2), 27,658 participants were analysed, including 7,795 from the pilot phase and 21,141 from the main phase; 1,353 (4.9%) had repeat assessments. The cohort comprised 49.9% women, and participants in the main phase were older than those in the pilot phase (mean age 72.3 vs 67.5 years; p<0.001). Median wear time was 14.0 days (IQR 11.9–14.0) in the pilot phase and 13.2 days (IQR 11.9–13.9) in the main phase, with 84.4% and 92.6% of recordings exceeding 7 days, respectively (Figure 3).

**Figure 2.**
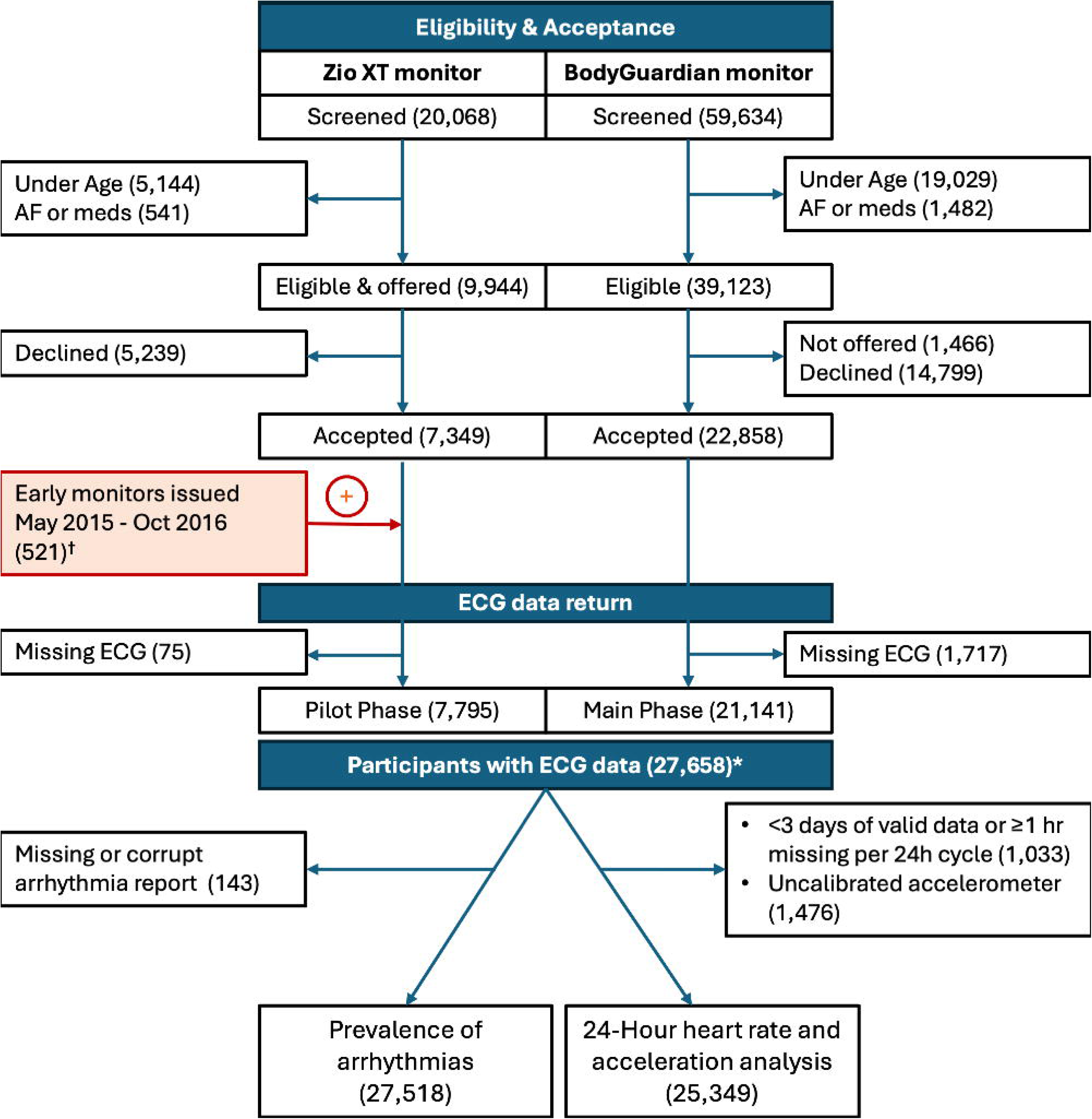
Exclusion diagram. Two analyses were conducted in this study: one assessing arrhythmia prevalence and the other examining the 24-hour heart rate profile. *\*1,353 participants took part in both study phases; ^†^Eligibility screening and acceptance rates were systematically recorded, except for the first 521 deployments; ^‡^Large number reflects data still awaiting processing at the time of writing*.

**Figure 3.**
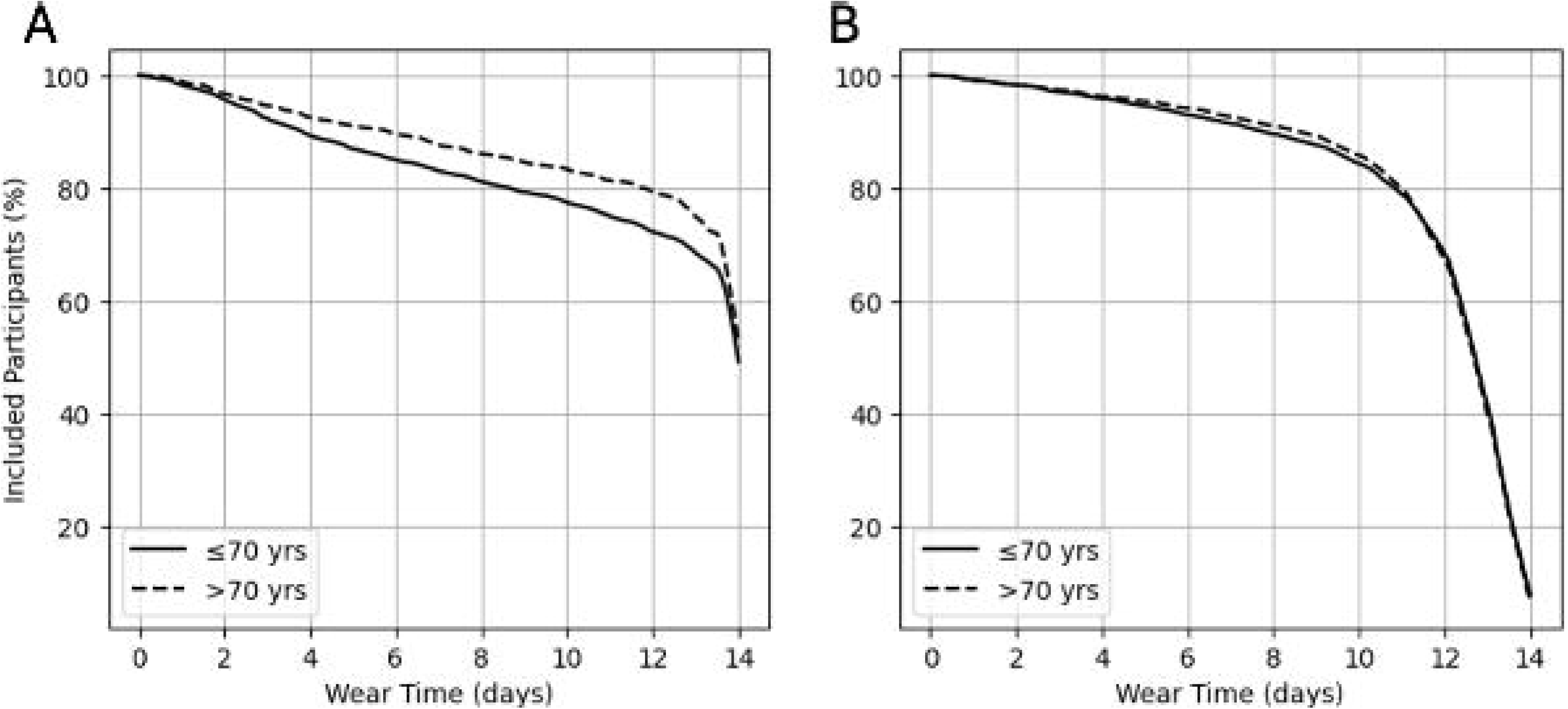
Monitor wear time in pilot (A) and main (B) phases, stratified by age in all participants with ECG data (n = 27,658).

Arrhythmia prevalence data were available for 27,581 participants (Figure 2). In the main study phase, 21,090 complete ECG quality control and arrhythmia reports from 21,015 participants were returned by MDT. Data quality was high, with analysable ECG covering most of the wear period (median 12.1 days [IQR 10.1–13.2]). At baseline, 51.5% of the participants were women and 59.2% were aged over 70 years. Several arrhythmias were detected, with age–sex differences in prevalence that were also evident in heart rate and physical activity (Figure 4; Supplementary Table 4). Subclinical AF occurred in 468 participants (2.4%), more frequent in men (3.2% vs 1.7%; p<0.001) and those >70 years (3.1% vs 1.4%; p<0.001). Most AF was paroxysmal (319; 68.1%); median AF burden 2.4% (IQR 0.8–6.3%). Median longest episode 326 min (IQR 109.5–728.5; max 5.7 days). The corresponding AF detection yield was 25.3% after 1 day, 50.2% after 3 days, and 74.3% after 7 days, with the remaining 25.7% detected during the second week (Supplementary Figure 1).

**Figure 4.**
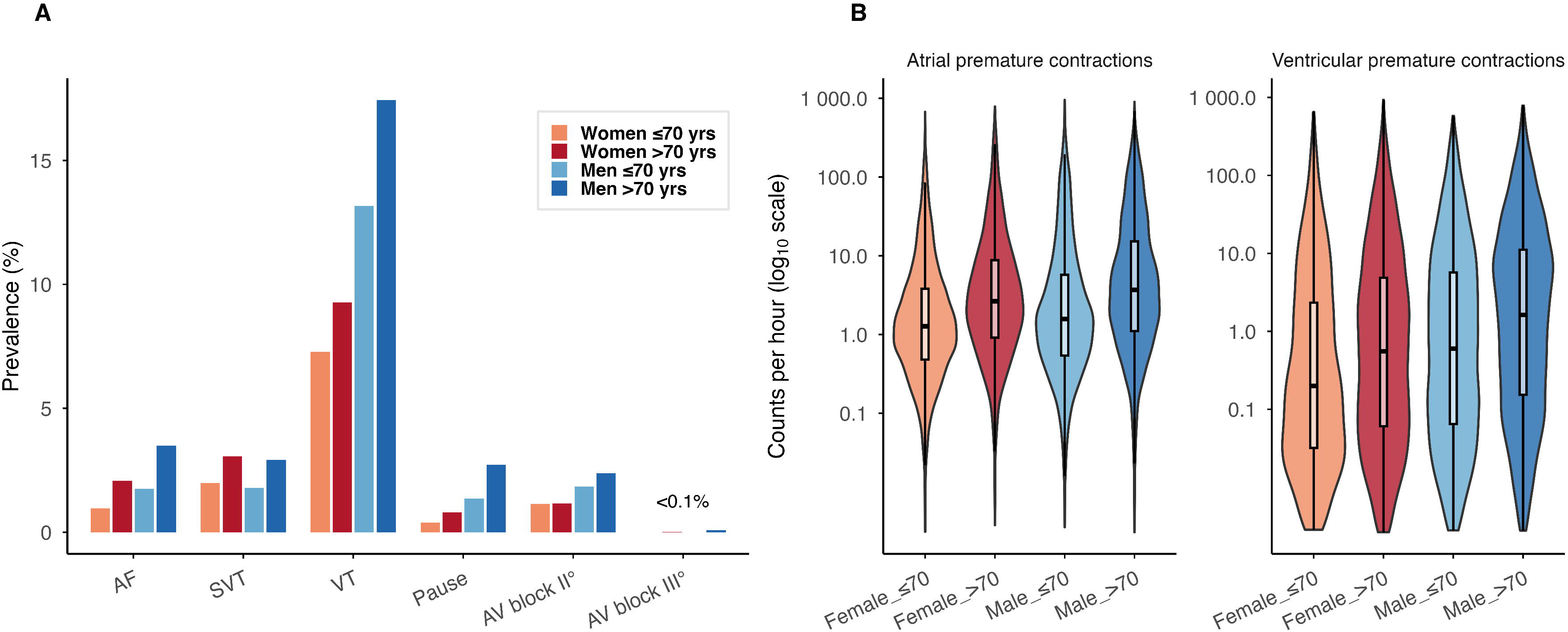
Age- and sex-related differences in arrhythmia prevalence and ectopy burden across four demographic groups.

SVT was observed in 537 participants (2.6%), with similar prevalence in women and men (2.6% vs 2.5%; p=0.7) but higher prevalence in older participants (3.0% vs 1.8%; p<0.001). The distribution of longest SVT episodes was strongly right-skewed, with 90.3% lasting less than one minute (Figure 5B). VT was detected in 2,549 participants (12.1%), more frequently in men than women (15.6% vs 8.4%; p<0.001) and in older compared with younger participants (13.6% vs 10.0%; p<0.001). Nearly all VT episodes (99.6%) were non-sustained, lasting less than 30 seconds (Figure 5C). Ectopic heart beats were detected in the vast majority of participants, with median hourly frequencies of 2.1 SVEs per hour (IQR 0.6–7.8) and 0.4 Ves per hour (IQR 0.4–5.1; Figure 4B; Supplementary Table 4). Ectopic burden was higher in men than women and in older than younger participants (all p<0.001).

**Figure 5.**
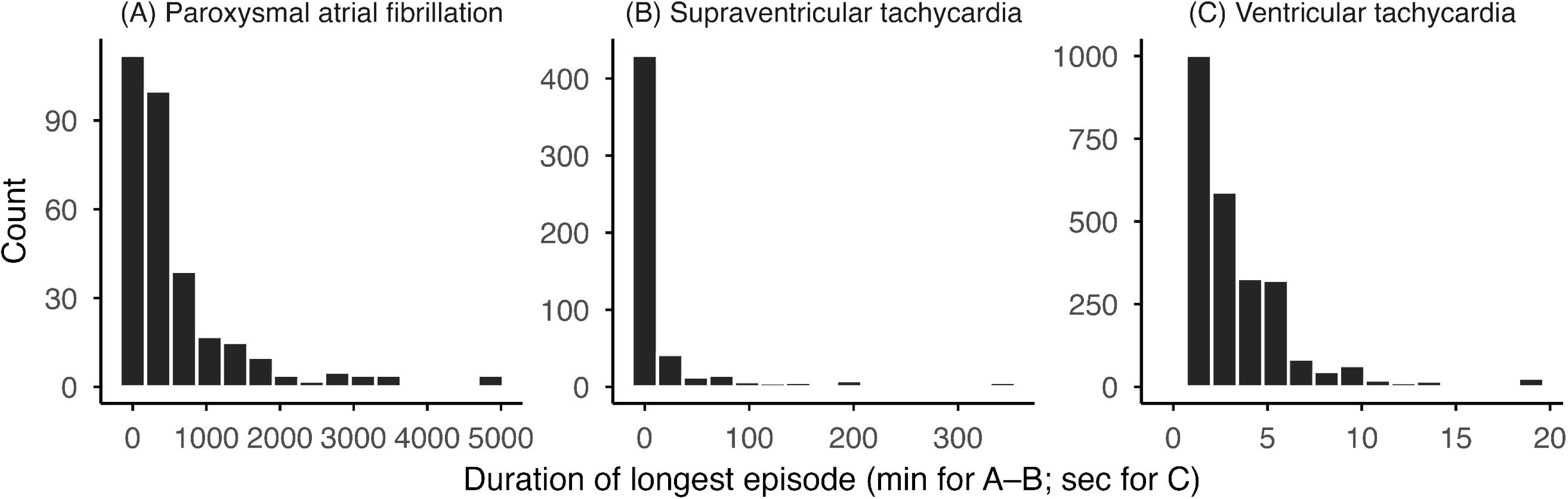
Distribution of the duration of the longest arrhythmia episode per participant for paroxysmal AF (A), SVT (B), and VT (C). Durations are shown in minutes for AF and supraventricular tachycardia (panels A and B) and in seconds for ventricular tachycardia (panel C). For plotting purposes, data were capped at the 99th percentile of episode duration.

Regarding conduction disorders, first-degree AV block was most prevalent (4,393 participants; 20.9%; Figure 4A), followed by second-degree (350; 1.7%) and third-degree AV block (7; <0.1%). AV block was more common in men than women (26.6% vs 15.9%; p<0.001) and in older than younger participants (23.9% vs 17.0%; p<0.001). Among participants with clinically significant AV block (second or third degree; 351 participants, 1.7%), 73 (20.8%) had their first episode detected more than 3 days after monitoring began.

An overview of arrhythmias observed in the pilot phase (Zio XT; 7,773 participants) is provided in Supplementary Table 5. ECG data quality was high, with most of the monitoring period analysable (median 13.7 days [IQR 11.5–14.0]). AF was observed in 130 participants (1.7%), with a detection yield similar to the main phase (78.7% by 7 days). Age and sex patterns were comparable, but SVT (6.2%) and VT (21.3%) were more prevalent, while sustained VT remained rare (0.1%).

We next examined the relationship between arrhythmia burden and physical activity. AF prevalence showed clear circadian variation, with higher occurrence at night, peaking around 3 am, and a morning nadir near 11 am (Figure 6A), coinciding with lower heart rate and physical activity (Figure 6C). In contrast, VE burden showed the opposite pattern, declining overnight to a trough around 5 am and increasing after waking, peaking at approximately 1 pm (Figure 6B), when heart rate and activity were highest (Figure 6C). Circadian variation in SVE burden was less pronounced, remaining within a narrow range of approximately 11.0 to 13.0 events per hour, with modest nocturnal predominance.

**Figure 6.**
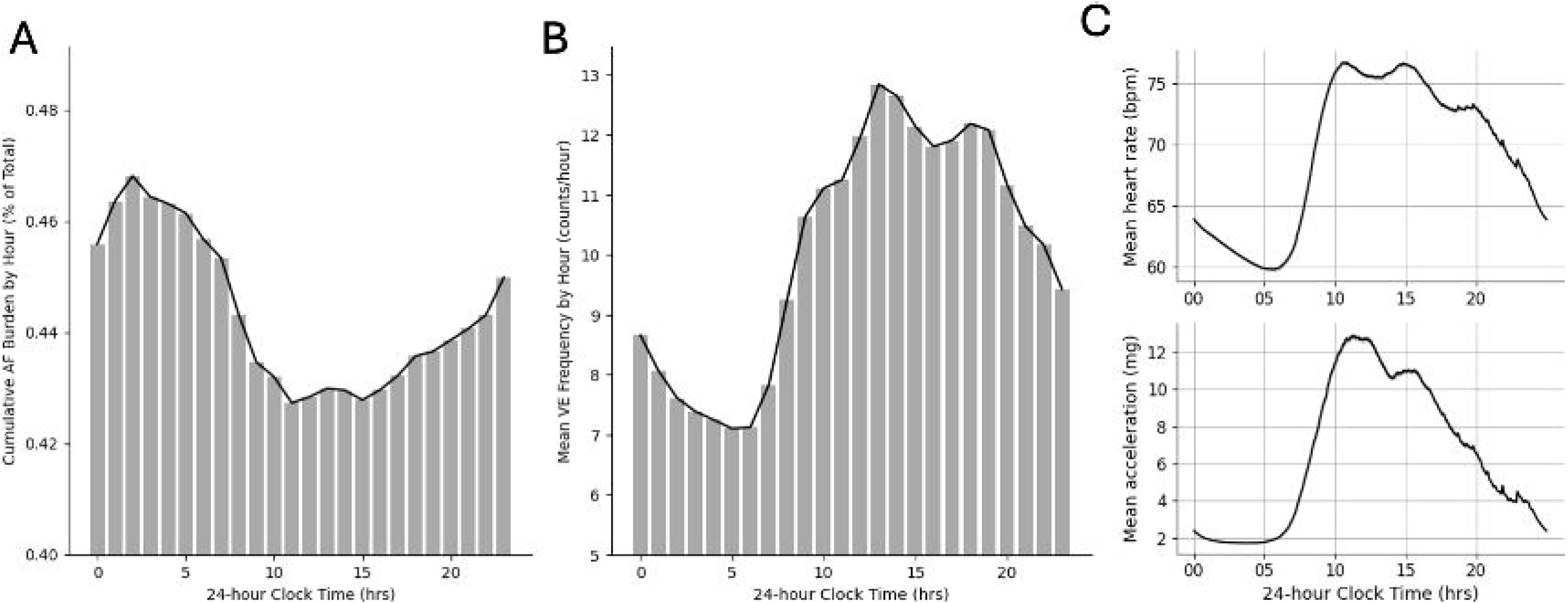
Diurnal variations in (A) cumulative burden of new atrial fibrillation (AF), (B) isolated ventricular ectopy (VE) counts, and (C) mean heart rate & acceleration. Data recorded during the main study phase in participants with at least 3 days of analysable data.

Longer-term variation in heart rate, signal quality, and physical activity patterns was assessed in 25,349 participants (91.1%) with at least 3 days of ECG data analysable by the in-house beat-to-beat heart rate detection pipeline (Figure 2). In this group, the median analysable ECG duration was 12.3 days (IQR 10.3–13.4), during which ECG quality varied diurnally, with higher quality at night and greater daytime noise, more pronounced in BodyGuardian MINI recordings (Supplementary Figure 2). Signal quality slightly declined over the 14-day monitoring period despite stable heart rate, which was not observed in Zio XT recordings (Supplementary Figure 3). Accelerometer data quality was high, with more than 99.9% analysable data; 27,895 datasets contained at least 3 analysable days, but 1,484 (5.5%) recordings could not be calibrated (Figure 2). Example heart rate and activity traces are shown in Supplementary Figure 4.

Mean heart rate and acceleration demonstrated clear 24-hour circadian rhythms in both study phases (Figure 6C; Supplementary Figures 5 and 6). Values increased after approximately 06:00, peaked between 09:00 and 10:00, and remained stable until mid-afternoon, then declined through the evening. Women had higher mean heart rates than men across the 24-hour period, but similar temporal profiles. Older participants showed lower daytime heart rate and acceleration than younger participants, with the largest differences in the afternoon. Heart rate profiles showed three recurrent peaks, in the morning (09:00–10:00), mid-afternoon (around 15:00), and early evening (around 19:00), mirrored by acceleration profiles.

A total of 1,345 participants had repeated arrhythmia assessments, with a median interval of 5.0 years (IQR 2.3–5.2) between recordings. Agreement in arrhythmia burden across assessments was moderate for AF (intraclass correlation coefficient [ICC] 0.58, 95% CI 0.55–0.62) and lower for SVE and VE (SVE: ICC 0.38, 0.34–0.43; VE: ICC 0.37, 0.33–0.42). Among 1,124 participants with at least 3 days of ECG and accelerometry at both assessments, use of a consistent device sequence (Zio XT followed by BodyGuard MINI) was associated with moderate-to-strong intra-individual correlations for mean heart rate and acceleration (Pearson r = 0.85 and 0.69, respectively). Twenty-four-hour heart rate and acceleration profiles showed strong within individual correlations across assessments, indicating stable circadian patterns over time (median r 0.82 [IQR 0.73–0.87] for heart rate and 0.63 [0.52–0.70] for acceleration; Supplementary Figure 7).

## Discussion

The UK Biobank cardiac monitoring study is the largest population-based dataset of prolonged ambulatory ECG monitoring to date, comprising nearly 28,000 participants, including repeat assessments in more than 1,300 individuals. It provides high-quality ECG and accelerometry data integrated with detailed information on the prevalence, burden, and temporal patterns of arrhythmias and conduction disorders, with enrolment expected to exceed 35,000 participants at completion.

Consistent with previous studies^3,17,18,26^, 14-day ECG monitoring showed that short term recordings substantially underestimate paroxysmal arrhythmia prevalence, highlighting the value of prolonged monitoring for population surveillance. Only about a quarter of paroxysmal atrial fibrillation episodes occurred within the first 24 hours, while more than a quarter were detected during the second week. The scale and duration of this dataset, an order of magnitude larger than prior population based cohorts with prolonged monitoring, enable more precise estimation of subclinical arrhythmia burden and investigation of temporal correlates.

A substantial burden of arrhythmias, including newly detected AF, was identified. Prevalence estimates derived from wearable ECG monitoring exceeded those from health records, reflecting the detection of asymptomatic and intermittent disease and highlighting a meaningful detection gap. Findings were consistent with prior population studies using similar monitoring approaches, including MESA (N=804), ARIC (N=2,244), and SAFARIS (N=527)^17,18,26^. Additionally, it confirmed established sex specific differences in arrhythmia prevalence.

Approximately half of SVT episodes in the main phase lasted one minute or less, whereas more than one-fifth exceeded ten minutes, indicating that sustained SVT may occur silently. Direct comparison with prior studies is complicated by differences in episode definition. SAFARIS^18^ used iRhythm-based analysis and defined SVT as episodes lasting more than 15s rather than 30s, likely contributing to their higher reported prevalence (12.3% vs 2.3%). Consistent with this, applying a >15s threshold to pilot-phase recordings processed by the same supplier yielded comparable prevalence (14.8%). However, even using the original 30 s definition, SVT prevalence remained higher in the pilot than in the main phase, suggesting an additional influence of vendor-specific arrhythmia detection algorithms and analytic pipelines.

These factors might also explain the higher prevalences of VT in SAFARIS (25%) and the pilot phase (21%). However, sustained VT remained rare in both study phases and indicated that such events can occur in community-dwelling populations. High-grade atrioventricular block was observed less frequently than in MESA^26^ and SAFARIS^18^ (1.6% vs ∼3%), potentially also reflecting differences in age distribution or detection criteria.

### Circadian variation in arrhythmias

Prolonged monitoring enables characterisation of temporal variation in arrhythmias across multiple 24-hour cycles. Distinct circadian profiles were observed: AF prevalence peaked around midnight and reached a nadir in the late morning, consistent with prior Holter-based findings by Yamashita et al.^27^, now demonstrated at scale in a population-based cohort. In contrast, VE and VT occurred predominantly during daytime hours, when sympathetic drive and myocardial workload are higher, in line with observations from smaller patient-based studies^28–30^. Interestingly, these patterns differed in their relationship with physical activity. AF was more common during periods of low activity, particularly at night, suggesting a role for increased vagal tone, whereas VE tracked with daytime activity, consistent with sympathetic activation or increased myocardial demand^28^. Although mechanisms remain uncertain, these data support future investigation of circadian arrhythmia patterns and autonomic biomarkers for identifying susceptibility and modifiable risk.

### Weartime & signal quality

Prolonged wearable monitoring was well tolerated, with over 90% of participants providing more than 7 days of analysable ECG data, supporting the feasibility of 14 day patch monitoring in population based studies and consistent with other large scale sensor deployments^17,18,26^. No substantial differences in wear time or analysable signal duration were observed between women and men. Although BodyGuardian MINI recordings seem to show greater signal degradation over time than Zio XT, likely reflecting electrode size or placement, heart rate and 24 hour activity rhythms remained highly stable within individuals, consistent with prior UK Biobank activity monitoring studies.studies^23^.

To our knowledge, this study also represents the largest population-based dataset with repeat wearable ECG measurements, providing a unique opportunity to examine the long-term stability of cardiac rhythm traits, including (circadian) heart rate measures and arrhythmia burden, at larger scale. Across repeated assessments separated by a median of five years, heart rate and physical activity measures showed equivalent reproducibility to the most reproducible imaging measurements (median≈85%)^31^ and the activity phenotypes exhibit equivalent reproducibility to wrist-worn accelerometer measurements^32^, supporting the robustness of wearable-derived physiological phenotypes. Agreement in arrhythmia burden was more modest. This might reflect the episodic nature of many rhythm disturbances, although at present interpretation of long-term repeatability is limited by the absence of data on medication use or other changes between assessments that may have influenced physiological measures and arrhythmia burden.

Several limitations should be acknowledged. First, the restriction to individuals aged 60 years or older limits generalisability to younger populations. Second, differences in sensor specifications, signal processing pipelines, and anatomical placement across devices (Zio XT over the left pectoral region vs BodyGuardian MINI at the mid sternum), together with arrhythmia analyses performed by different teams, may have introduced systematic differences in arrhythmia detection between study phases. Although preprocessing steps were applied to harmonise data, residual differences in accelerometry signals cannot be excluded, particularly given the low sampling frequency of the Zio XT device, which may limit detection of brief activity or fine grained movement. Finally, awareness of monitoring may have influenced physical activity or sleep behaviours during the recording period.

In conclusion, this is the largest population-based study of its kind. Extended ECG monitoring enabled the detection of subclinical arrhythmias and long-term physiological rhythms in older adults. Linkage to brain and cardiac imaging, multi-omics, and clinical outcomes in the UK Biobank will enable a detailed evaluation of the natural history of asymptomatic rhythm disturbances and their associations with brain health.

## Data Availability

All data produced in the present study will be made available in the future upon request to UK Biobank.

https://www.ukbiobank.ac.uk/

## Acknowledgments

This work was supported by the British Heart Foundation (RG/18/6/33576) and the Wellcome Trust (223100/Z/21/Z). Additional support was provided by a University of Oxford British Heart Foundation Centre of Research Excellence Fellowship awarded to SvD.

