## Supplemental Material for "Population-Wide Assessment of Heart Rhythm and Physical Activity from 14-Day Recordings: The UK Biobank Cardiac Monitoring Study"

**Supplemental Table 1 | Acceptance rates recorded during pilot phase;** Detailed demographic breakdowns (by sex and age) were only available from August 2016.

| | Acceptance by age & gender, all centres, to 13/12/2018 | | | | | | | | | | | |
|---|---|---|---|---|---|---|---|---|---|---|---|---|
| | >=60 <65 | | >=65 <70 | | >=70 <75 | | >=75 <80 | | >=80 | | All | |
| | Eligible | Accepted | Eligible | Accepted | Eligible | Accepted | Eligible | Accepted | Eligible | Accepted | Eligible | Accepted |
| **Female** | 2122 | 1224<br>57.7% | 2219 | 1189<br>53.6% | 1627 | 888<br>54.6% | 493 | 272<br>55.2% | 14 | 9<br>64.3% | 6475 | 3582<br>55.3% |
| **Male** | 1645 | 1025<br>62.3% | 1992 | 1198<br>60.1% | 1774 | 1110<br>62.6% | 680 | 417<br>61.3% | 22 | 17<br>77.3% | 6113 | 3767<br>61.6% |
| **Total** | 3767 | 2249<br>59.7% | 4211 | 2387<br>56.7% | 3401 | 1998<br>58.7% | 1173 | 689<br>58.7% | 36 | 26<br>72.2% | 12588 | 7349<br>58.4% |

**Supplemental Table 2 | Acceptance rates recorded during main phase**

| | Acceptance by age & sex | | | | | | | | | | | |
|---|---|---|---|---|---|---|---|---|---|---|---|---|
| | >=60 to <65 | | | >=65 to <70 | | | >=70 | | | All | | |
| | Eligible | Offered | Accepted | Eligible | Offered | Accepted | Eligible | Offered | Accepted | Eligible | Offered | Accepted |
| **Female** | 335 | 331<br>98.8% | 243<br>73.4% | 6400 | 6124<br>95.7% | 3819<br>62.4% | 13193 | 12690<br>96.2% | 7418<br>58.5% | 19928 | 19145<br>96.1% | 11480<br>60.0% |
| **Male** | 243 | 238<br>97.9% | 168<br>70.6% | 5256 | 5063<br>96.3% | 3233<br>63.9% | 13696 | 13211<br>96.5% | 7977<br>60.4% | 19195 | 18512<br>96.4% | 11378<br>61.5% |
| **Total** | 578 | 569<br>98.4% | 411<br>72.2% | 11656 | 11187<br>96.0% | 7052<br>63.0% | 26889 | 25901<br>96.3% | 15395<br>59.4% | 39123 | 37657<br>96.3% | 22858<br>60.7% |

**Supplemental Table 3 |** Definitions of cardiac arrhythmias analysed in pilot phase (2015-2018).

| Arrhythmia event | Definition |
|---|---|
| Pause | R-R interval > 2 seconds |
| AFib (Atrial Fibrillation) | Irregular polymorphic atrial activity with atrioventricular deficit (> 30s) |
| SVT (Supraventricular Tachycardia) | Regular or slightly irregular non-sinus tachycardia with a narrow QRS complex (episode > 30s) |
| VT (Ventricular Tachycardia) | Regular or slightly irregular tachycardia with a wide QRS complex with heart rate > 100 bpm (≥3 QRS complexes) |
| VF (Ventricular Fibrillation) | Irregular tachycardia with a wide polymorphic QRS complex |
| AV (Atrioventricular) Block I | Prolonged transmission from the atrial impulse to the ventricles with PR interval > 200ms |
| Isolated VE (Ventricular Ectopy) | Premature extra heartbeat originating from the ventricles, occurring earlier than the next expected normal beat |
| Isolated SVE (Supraventricular Ectopy) | Premature extra heartbeat originating from above the ventricles, occurring earlier than the next expected normal beat |
| VE Couplets / Triplets | Two or 3 VEs in a row |
| SVE Couplets / Triplets | Two or 3 SVEs in a row |
| Ventricular Bigeminy / Trigeminy | ≥Three cycles where every second (bigeminy) or third (trigeminy) beat is a ventricular ectopic |

**Supplemental Table 4** | Wear time, heart rate, and prevalence of arrhythmias in the main study phase as indicated by data supplier (Preventice BodyGuardian MINI monitor; 21,012 participants)

| Datasets, n | Women ≤70 yrs N = 4,544 | Women >70 yrs N = 6,043 | Men ≤70 yrs N = 3,761 | Men >70 yrs N = 6,667 |
|---|---|---|---|---|
| **Median wear time, days** | 13.2 (2.2) | 13.2 (2.2) | 13.2 (1.8) | 13.2 (1.8) |
| **Median analysable time, days** | 12.3 (3.0) | 12.3 (3.0) | 11.9 (3.1) | 11.9 (3.2) |
| **Mean heart rate, bpm** | 71.0 (9.0) | 70.6 (9.0) | 67.9 (11.0) | 66.9 (10.0) |
| **Mean acceleration, mg** | 6.8 (2.4) | 5.8 (2.1) | 7.3 (2.8) | 6.2 (2.3) |
| **Sinus pause, n** | 18 (0.4%) | 48 (0.8%) | 51 (1.4%) | 181 (2.7%) |
| **Atrioventricular block** | | | | |
| 1st degree, n | 590 (13%) | 1,064 (18%) | 794 (21%) | 1,945 (29%) |
| 2nd degree, n | 52 (1.1%) | 70 (1.2%) | 69 (1.8%) | 159 (2.4%) |
| 3rd degree, n | 0 (0%) | 1 (<0.1%) | 0 (0%) | 6 (<0.1%) |
| **Atrial premature contractions** | | | | |
| Median rate, counts/hour | 1.3 (3.3) | 2.5 (7.7) | 1.5 (5.0) | 3.2 (12.6) |
| Isolated, n | 4,492 (99%) | 5,906 (98%) | 3,678 (98%) | 6,301 (95%) |
| Couplets, n | 4,068 (90%) | 5,559 (92%) | 3,334 (89%) | 5,950 (89%) |
| Triplets, n | 3,458 (76%) | 4,960 (82%) | 2,757 (73%) | 5,220 (78%) |
| **Supraventricular arrhythmias** | | | | |
| Supraventricular tachycardia, n | 90 (2.0%) | 185 (3.1%) | 67 (1.8%) | 195 (2.9%) |
| Atrial flutter, n | 1 (<0.1%) | 3 (<0.1%) | 3 (<0.1%) | 18 (0.3%) |
| Atrial fibrillation (any), n | 44 (1.0%) | 125 (2.1%) | 66 (1.8%) | 233 (3.5%) |
| - *Paroxysmal, n* | 36 (0.8%) | 93 (1.5%) | 46 (1.2%) | 144 (2.2%) |
| - *Persistent, n* | 8 (0.2%) | 32 (0.5%) | 20 (0.5%) | 89 (1.3%) |
| **Ventricular premature contractions** | | | | |
| Median rate, counts/hour | 0.1 (1.6) | 0.3 (3.8) | 0.4 (4.7) | 1.3 (10.0) |
| Isolated, n | 3,987 (88%) | 5,427 (90%) | 3,448 (92%) | 6,326 (95%) |
| Couplets, n | 1,361 (30%) | 2,321 (38%) | 1,675 (45%) | 3,597 (54%) |
| Triplets, n | 369 (8.1%) | 661 (11%) | 586 (16%) | 1,442 (22%) |
| Bigeminy, n | 702 (15%) | 1,222 (20%) | 911 (24%) | 2,041 (31%) |
| Trigeminy, n | 769 (17%) | 1,328 (22%) | 869 (23%) | 2,119 (32%) |
| **Ventricular arrhythmias** | | | | |
| Idioventricular rhythm, n | 107 (2.4%) | 162 (2.7%) | 191 (5.1%) | 508 (7.6%) |
| Ventricular tachycardia, n | 331 (7.3%) | 560 (9.3%) | 495 (13%) | 1,163 (17%) |
| Ventricular fibrillation, n | 0 (0%) | 0 (0%) | 1 (<0.1%) | 0 (0%) |

*Persistent atrial fibrillation (AF) was defined as longest episode lasting >7 days. Median (IQR); Mean (SD); n (%)*

**Supplemental Table 5|** Wear time, heart rate, and prevalence of arrhythmias in pilot phase as indicated by data supplier (iRhythm Zio XT monitor; 7,773 datasets from 7,773 participants)

| Datasets, n | Women ≤70 yrs N = 2,821 | Women >70 yrs N = 963 | Men ≤70 yrs N = 2,699 | Men >70 yrs N = 1,290 |
|---|---|---|---|---|
| **Median wear time, days** | 14.0 (2.9) | 14.0 (1.1) | 14.0 (3.1) | 14.0 (0.8) |
| **Median analysable time, days** | 13.7 (3.2) | 13.7 (1.4) | 13.8 (3.3) | 13.8 (1.1) |
| **Mean heart rate, bpm** | 73.5 (9.0) | 72.2 (9.0) | 70.5 (11.0) | 68.8 (11.0) |
| **Mean acceleration, mg** | 8.0 (2.9) | 6.8 (2.3) | 8.4 (3.3) | 7.3 (2.7) |
| **Sinus pause, n** | 25 (0.9%) | 11 (1.1%) | 46 (1.7%) | 36 (2.8%) |
| **Atrioventricular block**, n | 58 (2.1%) | 12 (1.2%) | 47 (1.7%) | 37 (2.9%) |
| **Atrial premature contractions** | | | | |
| Median rate, counts/hour | 2.6 (4.6) | 4.3 (9.3) | 2.9 (5.9) | 5.2 (13.4) |
| Isolated, n | 2,819 (100%) | 960 (100%) | 2,683 (99%) | 1,280 (99%) |
| Couplets, n | 2,800 (99%) | 956 (99%) | 2,654 (98%) | 1,279 (99%) |
| Triplets, n | 2,053 (73%) | 795 (83%) | 1,867 (69%) | 1,059 (82%) |
| **Supraventricular arrhythmias** | | | | |
| Supraventricular tachycardia, n | 174 (6.2%) | 65 (6.7%) | 146 (5.4%) | 97 (7.5%) |
| Atrial fibrillation (any), n | 17 (0.6%) | 16 (1.7%) | 65 (2.4%) | 32 (2.5%) |
| - *Paroxysmal, n* | 17 (0.6%) | 14 (1.5%) | 53 (2.0%) | 24 (1.9%) |
| - *Persistent, n* | 0 (0%) | 2 (0.2%) | 12 (0.4%) | 8 (0.6%) |
| **Ventricular premature contractions** | | | | |
| Median rate, counts/hour | 1.5 (4.2) | 2.2 (7.6) | 1.7 (6.8) | 3.0 (12.5) |
| Isolated, n | 2,815 (100%) | 962 (100%) | 2,690 (100%) | 1,289 (100%) |
| Couplets, n | 2,230 (79%) | 816 (85%) | 2,102 (78%) | 1,093 (85%) |
| Triplets, n | 617 (22%) | 238 (25%) | 786 (29%) | 444 (34%) |
| Bigeminy, n | 634 (22%) | 284 (29%) | 802 (30%) | 521 (40%) |
| Trigeminy, n | 560 (20%) | 261 (27%) | 738 (27%) | 476 (37%) |
| **Ventricular arrhythmias** | | | | |
| Ventricular tachycardia, n | 373 (13%) | 185 (19%) | 670 (25%) | 430 (33%) |
| Ventricular fibrillation, n | 0 (0%) | 0 (0%) | 0 (0%) | 0 (0%) |

*Atrial fibrillation (AF) was considered persistent if total time spend in AF was >7 days (based on burden * analysable monitoring duration; episode duration was not available). Median (IQR); Mean (SD); n (%)*

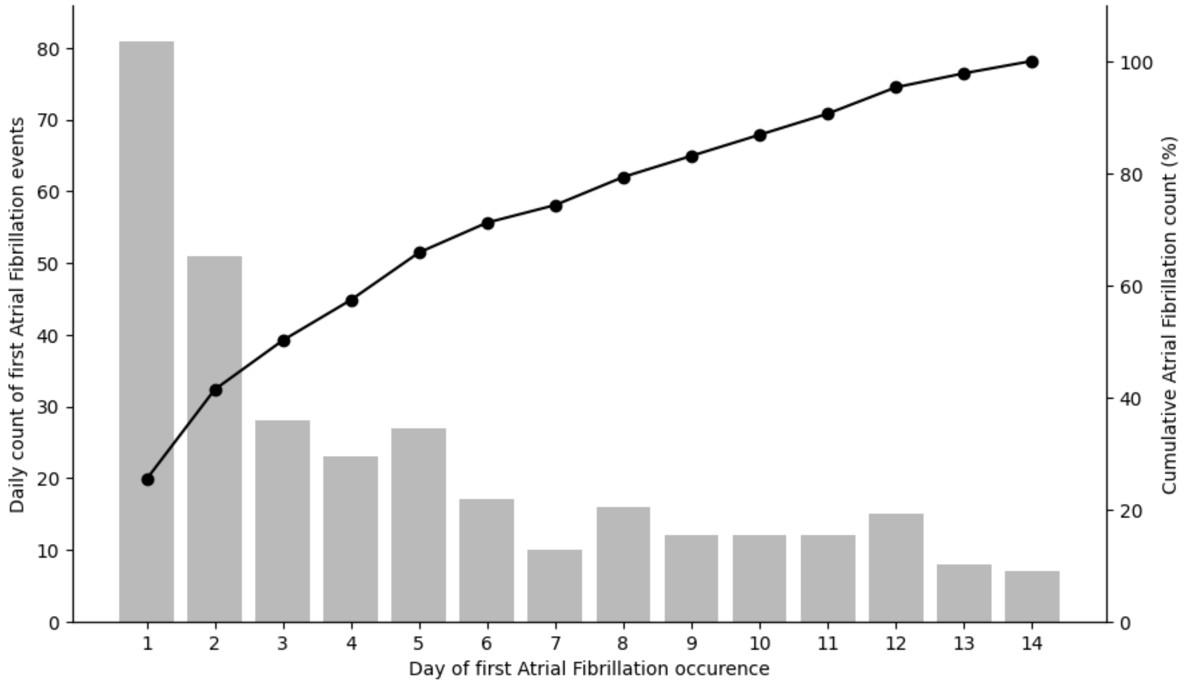


**Supplemental Figure 1 |** First occurrence of new atrial fibrillation in study phase 2 data.

**Supplemental Figure 2** | ECG signal quality indices as a function of the 24-hour cycle stratified by sex. Gray areas represent 95% confidence interval. Signal-to-noise ratios were only calculated in analysable ECG segments.

.

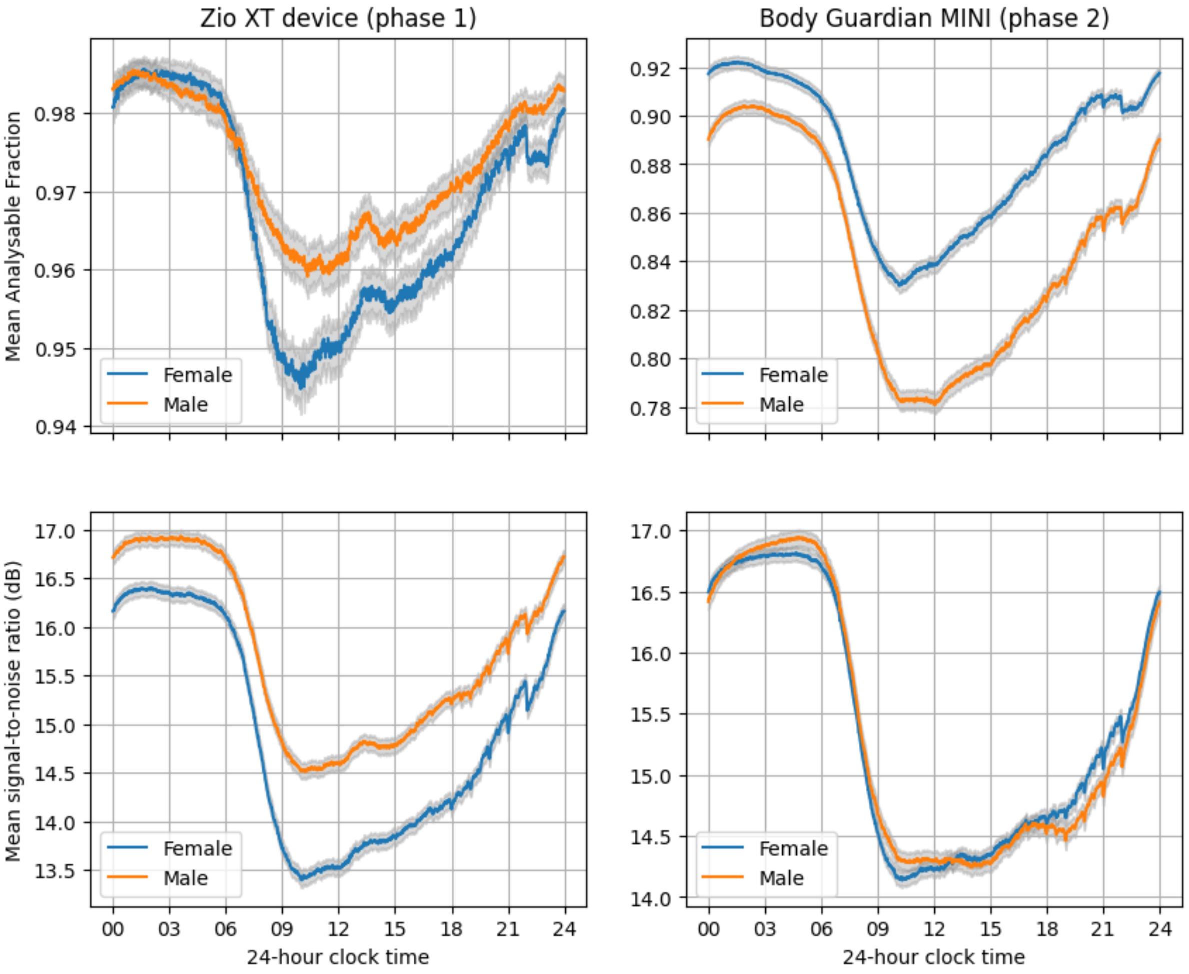

**Supplemental Figure 3** | ECG signal quality indices and mean heart rate plotted against 14-day monitoring period stratified by sex.

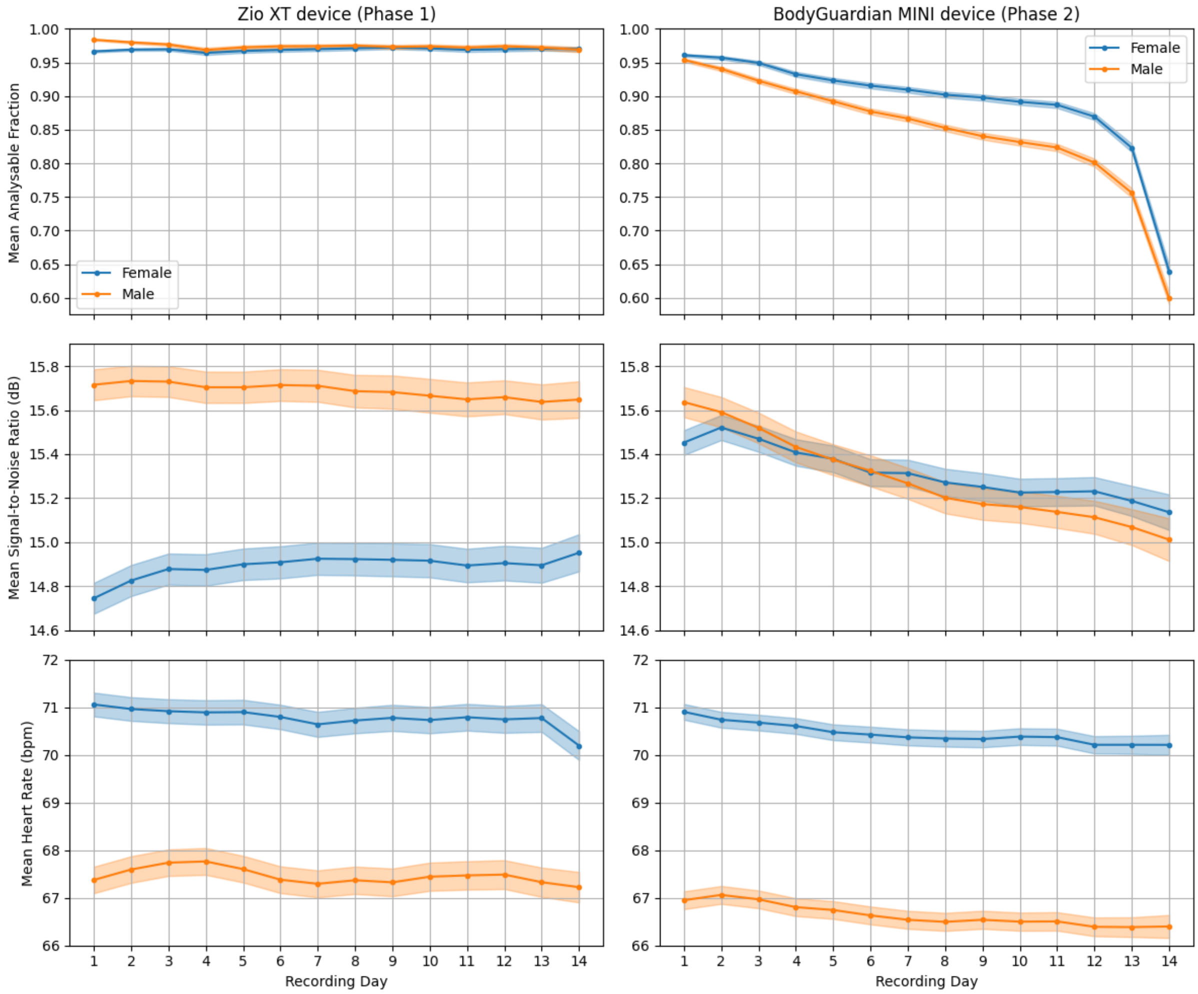

**Supplemental Figure 4** | Example recording with simultaneously plotted heart rate and acceleration data over 14 days of a single participant.

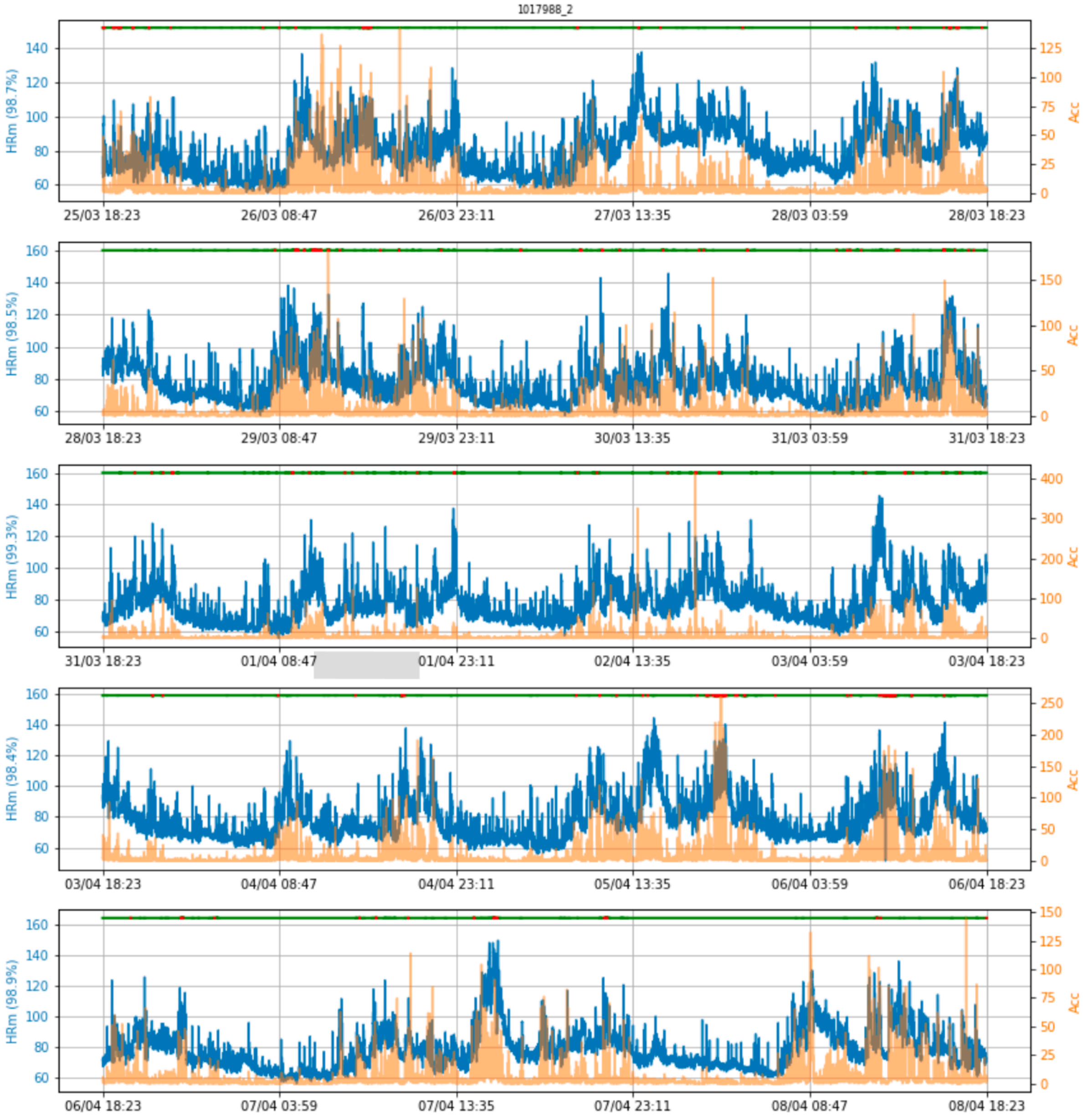

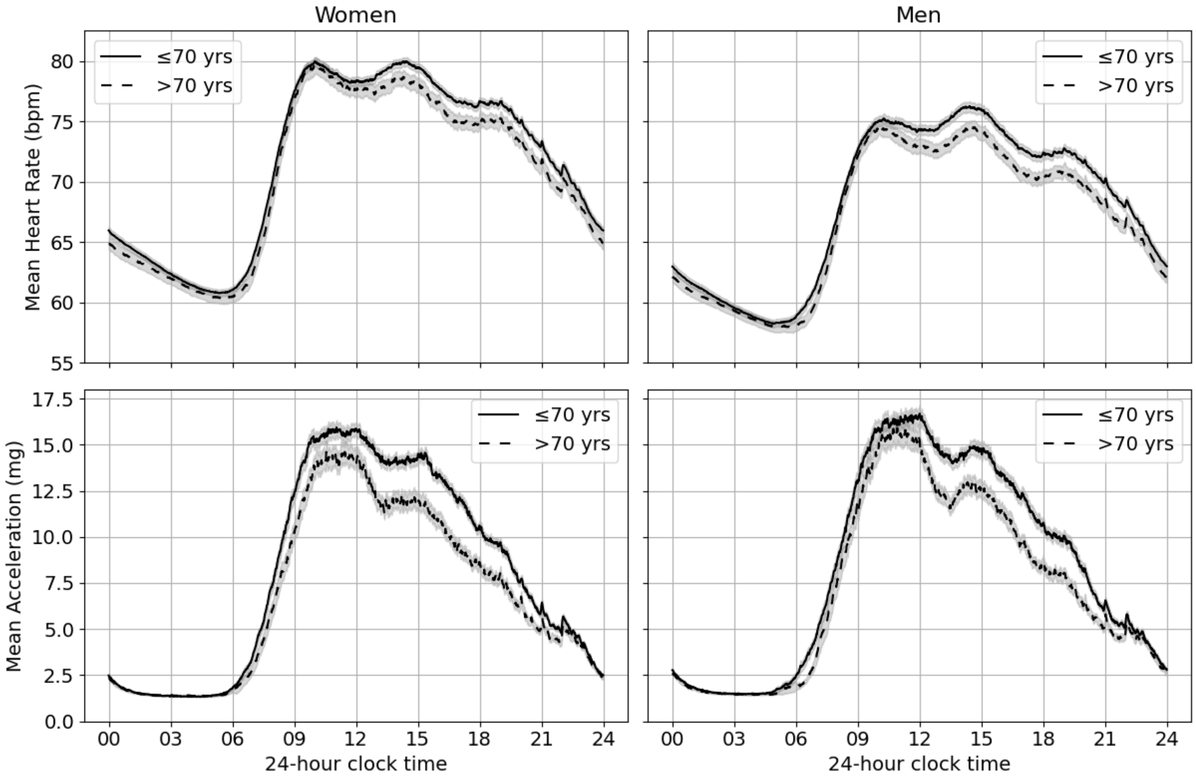


**Supplemental Figure 5** | Variation in mean heart rate and acceleration in pilot phase data across the day by sex and age. N=7,225 participants, shading bounds represent 95% confidence intervals.

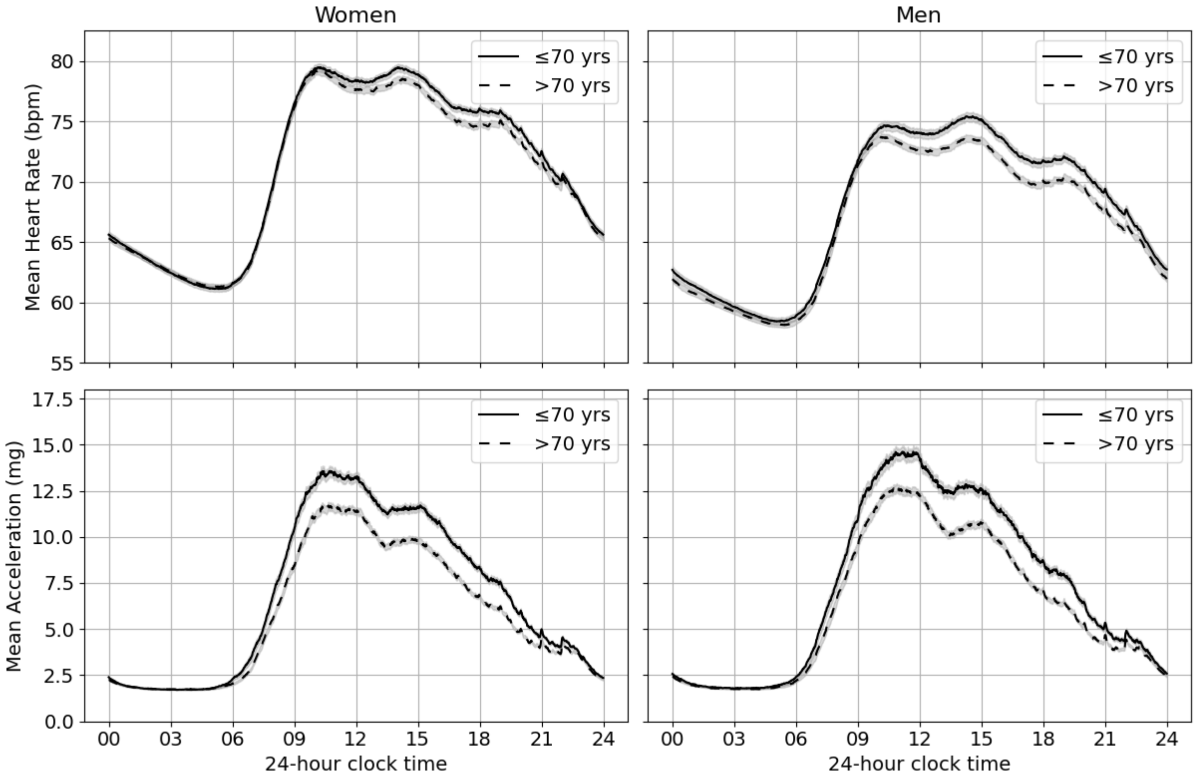


**Supplemental Figure 6** | Variation in mean heart rate and acceleration in main study phase data across the day by sex and age. N=19,248 participants, shading bounds represent 95% confidence intervals.

**Supplemental Figure 7** | Intra-individual correlation of heart rate and acceleration

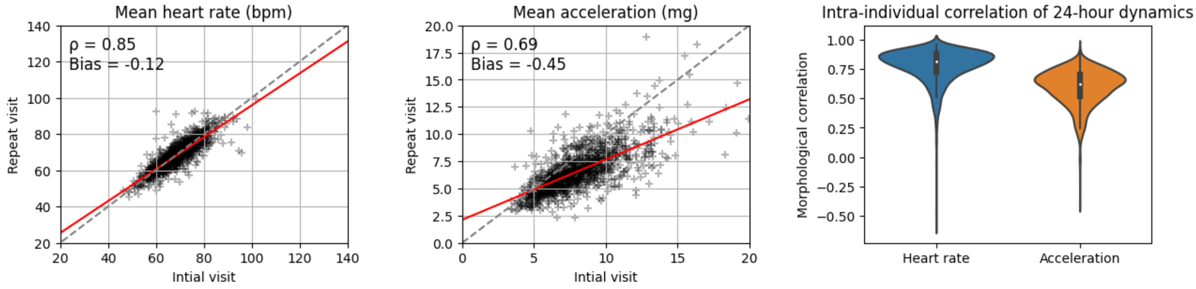


Analyses were performed in a subset of participants who had a repeat assessment (N=1,124). All initial recordings were obtained during pilot phase, and all repeat recordings during main phase.

**Supplemental Methods**

*Beat-to-beat RR interval extraction and ECG quality control*

Because manufacturer-processed data lacked beat-level information, independent beat-to-beat RR interval and ECG quality analyses were performed using a deep learning-based ECG segmentation algorithm for QRS detection. Training data comprised 75,000 manually annotated ten-second ECG segments from 5,000 randomly selected participants, sampled across diverse physical activity levels to reflect the variability of wearable ECG recordings. The algorithm estimated HR with a mean absolute error of 0.13 beats per minute (bpm) and mean squared error of 1.62 bpm on a 20% hold-out validation set. We used a heuristic approach to determine whether heart rate could be reliably estimated from each 10-s segment. Segments were excluded for non-wear (variance <0.001 mV$^2$), excessive noise (variance >2 mV$^2$), low amplitude (<25 µV), insufficient QRS complexes (<5), or unstable RR coverage (<75% of the segment or >1 RR outlier >1.8× the segment median).
